# Resources Required for Implementation of SARS-CoV-2 Screening in Massachusetts K-12 Public Schools in Winter/Spring 2021

**DOI:** 10.1101/2021.12.10.21267568

**Authors:** Stephanie S. Lee, Michelle Weitz, Kristin Ardlie, Amy Bantham, Michele Fronk Schuckel, Katey Goehringer, Caitlin Hogue, Rosy Hosking, Kathleen Mortimer, Alham Saadat, Jill Seaman-Chandler, Benjamin P. Linas, Andrea Ciaranello

## Abstract

**Importance:** CDC guidance emphasizes the importance of in-person education for students in grades kindergarten to 12 (K-12) during the COVID-19 pandemic. CDC encourage weekly SARS-CoV-2 testing of asymptomatic, unvaccinated students and staff (“screening”) to reduce infection risk and provide data about in-school SARS-CoV-2 prevalence where community incidence is high. The financial costs of screening assays have been described, but the human resource requirements at the school and district level to implement a SARS-CoV-2 screening program are not well known.

**Objective:** To quantify the resources required to implement a screening program in K-12 schools.

**Design, Setting, and Participants:** A consortium of Massachusetts public K-12 schools was formed to implement and evaluate a range of SARS-CoV-2 screening approaches. Participating districts were surveyed weekly about their programs, including: type of assay used, individual vs. pooled screening, approaches to return of results and deconvolution (identification of positive individual specimens) of positive pools, number and type of personnel implementing the screening program, and hours spent on program implementation.

**Main Outcomes and Measures:** Costs, resource utilization

**Results:** In 21 participating districts, over 21 weeks from January to June 2021, the positivity rate was 0.0%-0.21% among students and 0.0%-0.13% among educators/staff, and 4 out of 21 (19%) districts had at least one classroom transition to remote learning at any point due to a positive case. The average weekly cost to implement a screening program, including assay and personnel costs, was $17.00 per person tested; this was $46.68 for individual screenings and $15.61 for pooled screenings. The total weekly costs by district ranged from $1,644-$93,486, and districts screened between 58 and 3,675 people per week. The reported number of personnel working per week ranged from 1-5 to >50, and the total number of hours worked by all personnel ranged from 5-10 to >50.

**Conclusion and Relevance:** The human resources required to implement SARS-CoV-2 screening in Massachusetts public K-12 schools were substantial. Where screening is recommended for the 2021-22 school year due to high COVID-19 incidence (e.g., where vaccination uptake is low and/or more infectious variants predominate), understanding the human resources required to implement screening will assist districts policymakers in planning.

## INTRODUCTION

The US Centers for Disease Control and Prevention (CDC) encourages in-person learning in kindergarten through grade 12 (K-12) schools for students’ educational, physical, and emotional well-being.^1^ In 2020-21, the risks that students or educators/staff would acquire SARS-CoV-2 infection in a school setting were low when mitigation measures were well implemented;^2–4^ including masking, physical distancing, simple ventilation improvements, handwashing, diagnosis and contact tracing with appropriate isolation and quarantine, and vaccination once available.^5–7^ Some school communities also added weekly SARS-CoV-2 testing of asymptomatic people (“screening”). Screening may provide several benefits in K-12 schools.^8,9^ Where COVID-19 incidence is high, screening serves as an additional mitigation measure by identifying people with SARS-CoV-2 infection and isolating them before in-school transmission can occur. At all COVID-19 incidence levels, screening provides local, real-time information about SARS-CoV-2 prevalence in schools and may be reassuring to students, educators/staff, and their families.^5,10–15^

For the 2021-22 school year, CDC recommends screening of unvaccinated students and staff in communities where COVID-19 incidence is at least 10/100,000 people/week.^5^ Many K-12 schools are using substantially fewer mitigation measures than in 2020-21, including cohorting, universal masking, and physical distancing; at the same time, vaccination of al K-12 students is now available. Screening may play a role in monitoring the impact of both reduced mitigation measures and increased vaccination rates on in-school SARS-CoV-2 prevalence and transmission, especially as new and potentially more transmissible variants arise.^16,17^ Districts will need to weigh these potential benefits against the costs of screening programs. The costs of available SARS-CoV-2 screening assays vary widely, from <$5 to >$100 per person screened.^18,19^ While the cost of testing reagents and kits is well described, the additional costs associated with implementing a screening program are not well known.^20,21^ We sought to characterize the screening programs implemented in Massachusetts K-12 public schools and estimate the resources required for their implementation to inform future plans for funding and staffing of screening programs.

## METHODS

### Participating public school districts: STSS and DESE program

In August 2020, a consortium of 6 Massachusetts public K-12 schools, Safer Teachers Safer Students (STSS), was developed to implement and evaluate SARS-CoV-2 screening programs, negotiate lower assay costs, advocate for access to screening for all public school districts, and develop online resources.^14,22–25^ STSS grew to 33 districts by April 2021. Each STSS district initially contracted individually with chosen vendors; screening programs thus varied in assay type and cost, location of specimen collection, population screened, and screening frequency and schedule.

In January 2021, the Massachusetts Department of Elementary and Secondary Education (DESE) and Department of Public Health (DPH) offered pooled polymerase chain reaction (PCR)-based SARS-CoV-2 screening to all Massachusetts K-12 public schools providing in-person learning.^26^ The state provided test kits, support from testing service providers, and testing software. The program matched each participating district with a vendor from a state-approved list; assay type varied by vendor (PCR or next-generation sequencing [NGS]). Initially, all state-supported program vendors offered at-school pooling: students or staff collected anterior nares (AN) swabs and placed up to 10 swabs in a single collection tube. With at-school pooling, any pool reported as positive then required a second sample collection for “deconvolution” to identify the individual(s) from the original pool with SARS-CoV-2 infection. Most schools requested that members of a positive pool return to school for rapid antigen testing with a moderate-sensitivity and high-specificity assay provided by the state,^23^ or seek individual PCR testing at an outside facility (including free PCR testing offered at many state-sponsored testing sites).

Some STSS districts had previously contracted independently with vendors outside the state-supported program and continued to work with those vendors, while other STSS districts transitioned to the state-supported screening program.^27^ Reasons for continuing with previous vendors included familiarity with the vendor staff, consent processes, and result software. In addition, some vendors initially outside the state-supported program offered in-lab deconvolution, in which specimens from all individuals were retained and could be re-tested in the event of positive pool, without need for collection of a second sample. Over the course of Spring, 2021, the state-supported program expanded to include some vendors offering in-lab deconvolution.

### Data collection

We developed and administered surveys to STSS districts participating in any screening program (state-supported or independent). De-identified, aggregated data were entered in an online form weekly from January 18, 2021 or the week of screening implementation (whichever was later) to June 7, 2021. Survey questions included screening approaches: type of specimen (saliva or AN swab), type of assay (PCR or NGS), individual or pooled analysis, approach to result-return and deconvolution of positive pools, population screened (educators/staff, students, or subsets or combinations of these), and screening frequency (twice monthly, weekly, or twice weekly). Additional questions evaluated number eligible for screening, number participating in screening, number of positive pools, number testing positive, and resulting decisions, if any, about transitions to remote learning at the classroom, school, or district level. Questions about program implementation included: the type (role) of personnel involved, the number of people involved each week (in strata of 1-5, 5-10, 10-15, 15-20, 20-25, 25-50, and >50), and the number of hours spent by all personnel each week (strata of 5-10, 10-15, 15-20, 20-25, 25-50, and >50 hours). Cost questions included the cost to the district for assays and sample processing, inclusive of assay costs, laboratory fees, and shipping/courier fees. The full text of the survey questions is in Appendix A, and a publicly available dashboard, created by the STSS team, shows weekly participation and positivity rates from the survey responses (https://ma-k12testingcollaborative.org/).

We identified publicly available demographic and financial data for STSS districts participating in this study and for all Massachusetts public school districts, including student enrollment; number of staff employed; distribution of student gender and race/ethnicity; proportion of students who are economically disadvantaged, defined by participation in one or more state-administered programs (e.g., MassHealth or Supplemental Nutrition Assistance Program); proportions of students who are English language learners or students with disabilities, as defined by DESE;^28^ and total and in-district expenditures per pupil.^29^

### Costing approach

In general, we analyzed data about resource consumption in each district in a time updated manner, such that districts that adjusted their model over the course of the school year contributed data to the appropriate model of testing at each time point. We then multiplied those units of consumption by an estimate of the cost per unit to translate consumption to dollar outcomes.

### Costing labor

To estimate personnel costs, we made several simplifying assumptions necessitated by the structure of the available survey data. The surveys reported the number of people required for program implementation and the number of hours spent per week on implementation in strata (1-5, 5-10, etc.). We used the midpoint of each stratum in the base-case analysis; in sensitivity analyses, we used the lower and upper end of each stratum. We assumed that districts would only involve one of certain types of personnel; we assumed equal distribution of personnel type among all remaining types of personnel (e.g., school nurses, volunteers, etc.). We assumed that the total number of hours spent by all personnel was equally distributed among all involved personnel. For example, if data indicated that all staff in a district contributed 50 hours of labor, and there were 5 staff members, we assumed that each staff member contributed 10 hours. The sample calculations are in Appendix B.

We used data from the Bureau of Labor and Statistics to estimate wages and fringe benefits for the reported personnel types, and multiplied the estimated number of hours per week by hourly wages.^30–32^ We estimated the cost of parent and other volunteer time by using the average hourly wage in the United States.

### Costing Assays

We then calculated total (assay plus personnel) costs using data from each district’s most recent week of reporting. Due to the differences in assay cost for individual versus pooled screening, costs were calculated separately for each week depending on whether a district provided pooled or individual tests during that week. For weeks when pooled screeding was used, the cost of reflex testing (to deconvolute and identify which individual specimen(s) in a positive pool are positive) was estimated from the average number of positive pools and the average number of individuals included in a pool. We report the average per-person total assay cost (initial assay plus reflex testing). We then used the number of weeks in which either individual or pooled screening was offered to calculate a weighted average of the total weekly costs. We repeated these analyses varying several key assumptions: using the minimum and maximum of the reported range of the number of people involved in implementation and hours worked per week.

We calculated assay costs or total screening costs (including personnel costs) per person screened per week for all districts, as well as for those offering individual or pooled screening and those participating in the state-supported screening program or screening independently.

### Human subjects

This study was approved as “not human subjects research” by the Mass General Brigham institutional review board.

## RESULTS

### District characteristics

All 33 districts ever participating in STSS were invited to participate in this study, and 24 submitted data (response rate: 73%). Of these 24 districts, 21 (88% of responding, 64% of total) had both participation and costing data available and were included in the analysis. Characteristics of the 21 participating districts and of all Massachusetts public K-12 districts are shown in Table 1. The 21 districts represented approximately 10% of all schools, students, and teachers in the state; participating districts included a median of 8 schools, 3,597 students, and 270 teachers per district. Compared to the state overall, participating STSS districts included a greater proportion of White students (63.9% vs. 56.7%); similar proportions of Asian (7.4% vs. 7.2%), Native Hawaiian/Pacific Islander (0.0% vs. 0.1%), and mixed-race students (5.1% vs. 4.1%); and smaller proportions of Black and Hispanic students (11.0% vs. 31.6%), economically disadvantaged students (10.8% vs. 36.6%), and English language learners (4.4% vs. 10.5%). Participating districts’ publicly reported median in-district and total expenditures per pupil ($18,986 and $19,380) were higher than for the state overall ($16,588 and $17,150), as were average teacher salaries ($86,331 vs. $82,349).

**Table 1.** District Characteristics from 21 Participating Massachusetts K-12 School Districts

| ***District Characteristics*** | ***Districts***<br>***N=21*** | ***State***<br>***N= 400*** |
| --- | --- | --- |
| Student enrollment (all grades)<br>*Median (IQR)* | Total: 88,843 (9.7% of state)<br>Per district: 3,597 (2,019-5,891) | Total: 911,465<br>-- |
| Number of schools (N)<br>*Median (IQR)* | Total: 176 (9.6% of state)<br>Per district: 8 (4-11) | Total: 1,840<br>-- |
| Number of teachers (N)<br>*Median (IQR)* | Total: 7,508 (10.0% of state)<br>Per district: 270 (173-423) | Total: 75,146<br>-- |
| Number of districts participating in the state-funded program (N) | 12 | 190 (ever participating) |
| Enrollment by gender | *Median (IQR)* | (N) |
| Male | 1,810 (1,025-2,947) | 467,362 |
| Female | 1,754 (993-2,849) | 443,625 |
| Non-Binary | 1 (0-3) | 478 |
| Enrollment by race/ethnicity (%) | % (*IQR*) | (%) |
| Black | 4.4 (3.3-5.7) | 9.3 |
| Asian | 7.4 (3.4-14.9) | 7.2 |
| Hispanic | 6.6 (5.5-18.9) | 22.3 |
| Native American | 0.1 (0.0-0.2) | 0.2 |
| White | 63.9 (49.3-74.6) | 56.7 |
| Native Hawaiian, Pacific Islander | 0.0 (0.0-0.1) | 0.1 |
| Multi-race, Non-Hispanic | 5.1 (4.2-6.4) | 4.1 |
| Student Groups (%)** | % (*IQR*) | (%) |
| Economically disadvantaged | 10.8 (8.4-30.7) | 36.6 |
| English language learner | 4.4 (2.0-7.5) | 10.5 |
| Students with disabilities | 17.1 (16.0-18.8) | 18.7 |
| Finances (\$) | *Median (IQR)* | |
| In-District expenditures per pupil | \$18,986 (\$15,070-19,677)* | \$16,588* |
| Total expenditures per pupil | \$19,380 (\$16,049-20,825)* | \$17,150* |
| Teacher average salary (\$) | \$86,331 (80,382-92,906)* | \$82,349* |

\* Data are from the most recent available year
\*\* As defined by the Massachusetts Department of Elementary and Secondary Education<sup>28</sup>

### Characteristics of screening programs

Of the 21 districts, at the time of the most recent reporting, 21 (100%) screened both educators/staff and students (Table 2). The majority (67%) screened students at all grade levels. More districts screened weekly (20) than twice monthly (1), and more districts used pooled screening (20) than individual screening (1). At the most recent reporting, twelve districts (57%) participated in the state-supported screening program. Over the entire study period, 3 (14%) districts reported using individual screening for a total of 12 district-weeks. All districts reported using pooled screening at some point during the study period, for a total of 178 district-weeks. During the last reporting week, 1 district used individual PCR screening to screen educators/staff and pooled screening for students, while 20 districts reported using pooled screening for both educators/staff and students.

**Table 2.** Characteristics of school-based SARS-CoV-2 surveillance programs in participating districts

| Characteristic | Most recent reporting<br>(n= 21 districts)<br>N (%) |  |
| --- | --- | --- |
| Target population for surveillance testing*<br>Educators, staff, and students | 21 (100%) |  |
| Frequency of screening<br>Weekly<br>Twice monthly | 20 (95%)<br>1 (5%) |  |
| Grades of students included in the program*<br>All grades<br>Elementary only<br>Elementary, Middle only<br>Middle, High only<br>High only | 14 (67%)<br>1 (5%)<br>2 (9%)<br>2 (9%)<br>2 (9%) |  |
| Screening strategy (educators/staff)*<br>Individual<br>Pooled | 1 (5%)<br>20 (95%) |  |
| Screening strategy (students)*<br>Pooled | 21 (100%) |  |
|  | All districts | Per district<br>(Median, IQR) |
| N of students and educators/staff ever screened | 271,246 | -- |
| N screened (per week)*<br>Educators/staff<br>Students | 4,923<br>20,991 | 216 (109, 354)<br>781 (606, 1,162) |
| % participating among those offered screening*<br>Educators/staff<br>Students | 40%<br>38% | 41% (35%, 76%)<br>43% (28%, 63%) |

\*Data are from each district's most recent reporting week during the study period.

### Outcomes of screening programs

Educators/staff and students underwent a total of 271,246 tests. In the first week of the study period, 5,168 students from 5 districts (1,034 students per participating district) were offered screening, and 3,424 students from 5 districts (685 students per participating district) underwent screening (Figure 1). These numbers increased to 4,137 students/district offered screening and 1,454 students/district screened in week 21. For educators and staff, in week 1,763 educators/staff per district were offered screening and 443 staff/ district were screened; in week 21,952 educators/staff per district were offered screening and 347 educators/staff per district were screened. While the absolute number changed over the study period, so did the number of districts reporting. The participation rates among all educators and/or staff (Figure 1, panel A) remained fairly constant, while participation among all students declined (Panel B). The positivity rate among students was 0.0%-0.21% and 0.0%-0.13% among educators/staff. Among the 21 school districts, 4 (19%) reported a classroom, school, or district temporarily transitioning to remote learning due to a case identified through the screening programs.

**Figure 1.**
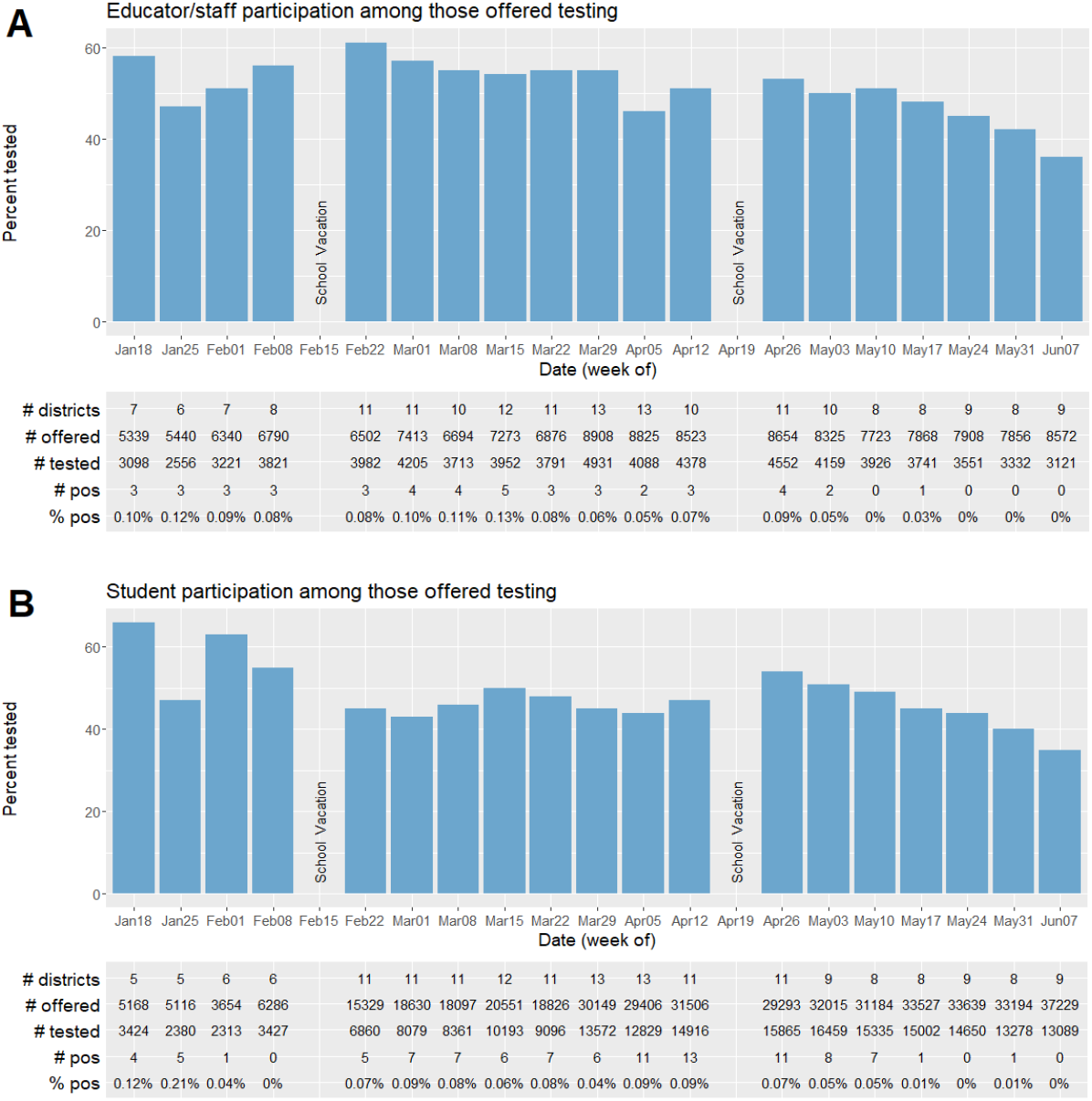
Massachusetts School-Based SARS-C0V-2 Testing Volume Per Week 1/18/2021-6/7/2021 The bar graphs represent the educators/staff and students tested among those offered testing (%) from Jan 18 to Jun 07. The columns underneath the bar graphs show the number of districts, number offered testing, number tested, number of those who tested positive, and the positivity rate (%). The positivity rate ranged from 0.0%-0.13% for educators and staff and 0.0%-0.21% for students.

### Resource utilization and costs associated with screening programs

Among the 21 districts, the reported number and types of implementing personnel and personnel-hours spent in program implementation are shown in Table 3 for all districts, as well as for district-weeks of individual and pooled PCR screening and of state-supported and non-state-supported programs. Per person screened each week, average assay costs (including shipping and laboratory processing) were $12.60, average personnel costs were $4.27, and average total costs (assay plus personnel) were $17.00. In sensitivity analyses using the upper and lower bounds of reported personnel and time strata, average weekly per-person cost varied from $15.67 to $18.34.

**Table 3.** Resource Utilization and Costs Associated with Implementing Screening Programs

| **Resource Utilization<sup>1</sup>** |  |  |  |  |  |  |  |
| --- | --- | --- | --- | --- | --- | --- | --- |
| *N (%)* | *All districts*<br>*n=21* | *Individual*<br>*screening*<br>*(educators/*<br>*staff only)*<br>*n=1* | *Pooled*<br>*screening*<br>*n=20* | *Pooled*<br>*screening,*<br>*deconvolute*<br>*in-lab*<br>*n=8* | *Pooled*<br>*screening,*<br>*deconvolute*<br>*in-school*<br>*n=13* | *State-supported*<br>*screening*<br>*program*<br>*n=12* | *Non-state-supported*<br>*screening program*<br>*n=9* |
| Implementation personnel |  |  |  |  |  |  |  |
| Town or city staff | 3 (14%) | 0 (0%) | 3 (16%) | 2 (25%) | 1 (8%) | 0 (0%) | 3 (33%) |
| District Physician | 2 (10%) | 0 (0%) | 2 (11%) | 1 (13%) | 1 (8%) | 1 (8%) | 1 (11%) |
| Educators/staff | 10 (48%) | 1 (100%) | 9 (47%) | 4 (50%) | 6 (46%) | 5 (42%) | 5 (55%) |
| Superintendent | 2 (10%) | 0 (0%) | 2 (10%) | 0 (0%) | 2 (15%) | 2 (17%) | 0 (0%) |
| Paid project managers | 6 (29%) | 0 (0%) | 6 (32%) | 2 (25%) | 4 (31%) | 4 (33%) | 2 (22%) |
| School nurses at each school | 21 (100%) | 1 (100%) | 20 (100%) | 8 (100%) | 13 (100%) | 12 (100%) | 9 (100%) |
| School nurses at each district | 13 (62%) | 1 (100%) | 12 (63%) | 7 (88%) | 6 (46%) | 6 (50%) | 7 (78%) |
| District health department | 3 (14%) | 0 (0%) | 3 (16%) | 1 (13%) | 2 (15%) | 2 (17%) | 1 (11%) |
| Parent/other volunteers | 7 (33%) | 0 (0%) | 7 (37%) | 3 (38%) | 4 (31%) | 3 (25%) | 4 (44%) |
| Number of people involved in implementation |  |  |  |  |  |  |  |
| 1-5 | 3 (14%) | 0 (0%) | 3 (15%) | 0 (0%) | 3 (23%) | 3 (25%) | 0 (0%) |
| 5-10 | 5 (24%) | 1 (100%) | 4 (20%) | 0 (0%) | 5 (38%) | 5 (42%) | 0 (0%) |
| 10-15 | 4 (19%) | 0 (0%) | 4 (20%) | 1 (13%) | 3 (23%) | 3 (25%) | 1 (11%) |
| 15-20 | 3 (14%) | 0 (0%) | 3 (15%) | 2 (25%) | 1 (8%) | 0 (0%) | 3 (33%) |
| 20-25 | 2 (10%) | 0 (0%) | 2 (10%) | 2 (25%) | 0 (0%) | 0 (0%) | 2 (22%) |
| >25 | 3 (14%) | 0 (0%) | 3 (15%) | 3 (38%) | 0 (0%) | 0 (0%) | 3 (33%) |
| >50 | 1 (5%) | 0 (0%) | 1 (5%) | 0 (0%) | 1 (8%) | 1 (8%) | 0 (0%) |
| Number of hours spent by personnel on implementation |  |  |  |  |  |  |  |
| 5-10 hours/week | 3 (14%) | 0 (0%) | 3 (15%) | 0 (0%) | 3 (23%) | 3 (25%) | 0 (0%) |
| 10-15 hours/week | 0 (0%) | 0 (0%) | 0 (0%) | 0 (0%) | 0 (0%) | 0 (0%) | 0 (0%) |
| 15-20 hours/week | 2 (10%) | 0 (0%) | 2 (10%) | 0 (0%) | 2 (15%) | 1 (8%) | 1 (11%) |
| 20-25 hours/week | 3 (14%) | 0 (0%) | 3 (15%) | 2 (25%) | 1 (8%) | 1 (8%) | 2 (22%) |
| >25 hours/week | 7 (33%) | 1 (100%) | 6 (30%) | 3 (38%) | 4 (31%) | 5 (42%) | 2 (22%) |
| >50 hours/week | 6 (29%) | 0 (0%) | 6 (30%) | 3 (38%) | 3 (23%) | 2 (17%) | 4 (44%) |

**Table 3.** Resource Utilization and Costs Associated with Implementing Screening Programs, continued
| Costs per person tested per week <sup>2</sup> |  |  |  |  |  |  |  |
| --- | --- | --- | --- | --- | --- | --- | --- |
|  | *All districts* | *Individual screening* | *Pooled screening* | *Pooled screening, deconvolute in-lab* | *Pooled screening, deconvolute in-school* | *State-supported screening program* | *Non-state-supported screening program* |
| Mean assay cost<sup>3</sup><br>SD, range | \$12.73<br>7.92, 5.00-31.54 | \$44.44<br>9.11, 34.33-52.00 | \$11.35<br>6.14, 5.00-25.00 | \$15.81<br>5.61, 6.98-25.00 | \$8.59<br>4.78, 5.00-18.32 | \$7.11<br>3.24, 5.00-15.04 | \$16.98<br>4.14, 12.18-25.00 |
| Mean personnel cost<sup>4</sup><br>SD, range | \$4.27<br>5.53, 0.36-23.34 | \$2.24<br>1.04, 1.27-3.34 | \$4.26<br>5.53, 0.36-23.34 | \$3.11<br>3.45, 0.61-11.33 | \$4.98<br>6.53, 0.36-23.34 | \$5.84<br>6.91, 0.36-23.34 | \$2.17<br>1.48, 0.61-5.20 |
| (Sensitivity analysis<sup>5</sup>) | (\$2.94, \$5.61) | (\$1.48, \$2.94) | (\$2.93, \$5.61) | (\$1.97, \$4.25) | (\$3.51, \$6.45) | (\$4.16, \$7.50) | (\$1.29, \$3.09) |
| Mean total cost<sup>6</sup><br>SD, range | \$17.00<br>7.75, 5.37-32.84 | \$46.68<br>8.08, 37.67-53.27 | \$15.61<br>6.54, 5.37-28.35 | \$18.92<br>4.76, 13.14-27.55 | \$13.57<br>6.80, 5.37-28.35 | \$12.95<br>6.52, 5.37-28.35 | \$19.15<br>4.86, 13.14-27.55 |
| (Sensitivity analysis<sup>5</sup>) | (\$15.67, \$18.34) | (\$45.92, \$47.38) | (\$14.28, \$16.96) | (\$17.78, \$20.06) | (\$12.10, \$15.04) | (\$11.27, \$14.61) | (\$18.27, \$20.07) |

1. Data are from each district's most recent reporting week during the study period. 2. Costs were calculated over a period of time rather than the most recent reporting 3. For districts that did not report the cost of assays, we estimated those costs based on the vendor used. 4. Estimated personnel costs were calculated from mean number of hours \* average publicly reported salary for each type of employee, weighted across types of employee reported. Numbers of personnel and hours were reported in strata; we assumed an inclusive upper bound for each stratum. For ranges where an upper bound was not explicitly specified (>25 and >50), we assumed a range of 25-50 and 50-150, respectively. 5. Sensitivity analyses: lower and upper bound of mean cost estimate, using lower and upper bounds of strata of number of personnel and personnel-hours spent. 6. Sum of reported assay and other costs plus estimated personnel costs

Average weekly per-person total cost was $46.68 for districts when using individual screening and $15.61 for districts when using pooled screening; the difference was due primarily to higher assay costs ($44.44 vs. $11.21). On average per person screened each week, non-state-supported programs compared to state-supported programs had higher assay costs ($16.98 vs. $7.11), lower personnel costs ($2.17 vs. $5.84), and higher total costs ($19.15 vs. $12.95). The main driver of costs across multiple strategies was the assay costs, which comprised 75% of costs for all districts, 73% of costs for district-weeks of pooled screening, 95% of costs for district-weeks of individual screening, 55% of costs for state-supported screening programs, and 89% of costs for non-state-supported screening programs (Figure 2).

**Figure 2.**
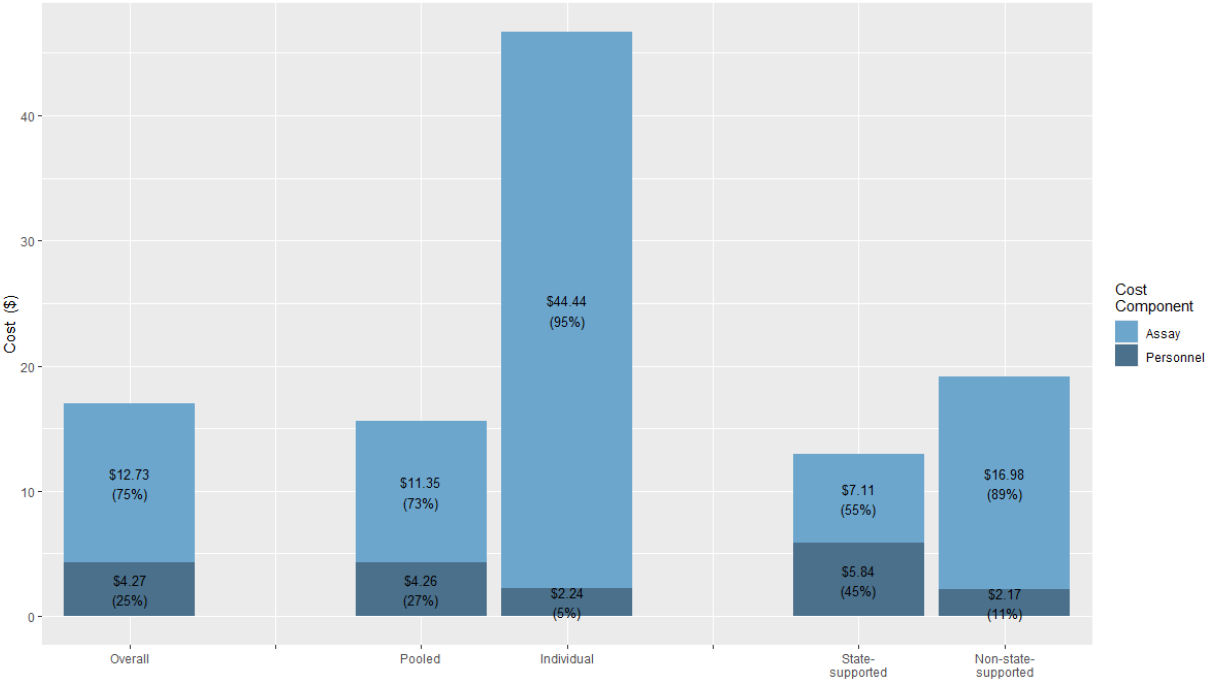
Components of average weekly cost per person tested The bar graphs represent the mean costs ($) for assay type (includes cost of initial assay as well as reflex testing, if applicable) and personnel and its percentage (%) in proportion to the total costs. Pooled and individual refer to the type of test; state-supported or non-state-supported refers to the source of funding for the screening programs. Across all the groups, assay type accounted for more of the costs, though with a greater proportion in the individual testing and for non-state-supported screening programs.

Of the 21 districts, 8 (38%) used in-lab deconvolution for positive pools and 13 (62%) used in-school deconvolution for positive pools. Compared to in-school deconvolution, in-lab deconvolution had higher average weekly per-person assay costs ($15.81 vs. $8.59), lower personnel costs ($3.11 vs. $4.98), and higher total cost ($18.92 vs. $13.57). Of the 13 districts using in-school deconvolution, 7 (54%) reported using Abbot SARS CoV-2 Binax NOW (tm), 3 (23%) reported using PCR, and 3 (23%) reported using both Abbot SARS CoV-2 Binax NOW (tm) and PCR. Compared to Abbot SARS CoV-2 Binax NOW (tm), PCR deconvolution led to higher weekly per-person average assay cost ($13.13 vs. $6.59), lower personnel cost ($2.45 vs. $6.57), and higher total costs ($15.58 vs. $13.34; not shown in Table 3).

## DISCUSSION

We estimated the cost of asymptomatic SARS-CoV-2 screening among students and staff in 21 Massachusetts public school districts, with two main findings. First, the cost of screening was approximately $17.00 per person per week when including assay and personnel costs, and the total cost was higher for individual than pooled screening ($46.68 vs. $15.61); 95% of the total cost for programs offering individual tests was due to the costs of the assays themselves. We noted wide variation in costs between districts, mainly driven by the selection of pooled vs. individual screening.

Second, we found that a wide-ranging and often large number of personnel (3-100 per district) and person-hours per week (8-100 per district) were required to implement screening programs. The type of personnel also varied by district, as some dedicated 5-6 full-time equivalents, while others relied on 7-8 part-time volunteers; we assigned the average US hourly wage for the cost of volunteer time. Additionally, the total number of hours spent by staff/volunteers on implementation ranged from 8 to 100 hours per week, although the majority spent more than 25 hours. Importantly, these hours were added to the time that school staff spent on additional COVID-19 mitigation strategies, such as contact tracing and redesigning schedules and facilities to support smaller class sizes and greater distancing; time for these activities was not reported in the surveys.

Assay costs reported by districts in this pilot study differ from others previously reported, partly due to prices negotiated between districts and vendors; other sources report higher individual testing costs ($50-$200) and lower pooled tests ($50.00 per pool or $5.00 per individual swab).^18,33,34^ In addition, the costs for pooled tests depend on the pool size; variation in pool sizes in our study may partly explain the per-person cost.^35^ Our estimated weekly personnel cost per person tested ($4.27) was not directly comparable to other reports ($99-198), which included personnel costs for additional mitigation measures, such as sanitation.^36^

Massachusetts was one of the first states to implement state-wide pooled screenings in K-12 school settings, thus serving as a model of the feasibility and advantages of the approach.^37,38^ While our analysis is limited to Massachusetts, these data provide useful information for schools across the U.S. For example, several schools in Washington use rapid antigen tests for assurance testing,^39,40^ Davis County in Utah conducts widespread screening when cases in a school exceed 1% of the school population,^41,42^ and Michigan’s health department launched a weekly COVID testing program solely for educators.^43,44^ Most states offer voluntary screening, while other states, such as New York, implemented a mandatory state-wide surveillance program, screening a sample of educators/staff and students in schools in areas at higher risk of COVID-19 transmission.^45,46^ While the cost per person tested will differ across settings, based on local labor markets and the cost of assays and reagents, this analysis provides an estimate of both the resources needed to implement testing and the key drivers of cost. We anticipate that the high-level findings – that testing is expensive and that the choice of testing model has a large impact on cost considerations – will likely be generalizable to most K-12 school settings.

As schools weigh the trade-offs of implementing a screening program, they must consider a range of benefits and costs. In our study, despite relatively high participation rates among students and staff during a period of high community incidence before vaccination, the screening program identified only a small number of SARS-CoV-2 infections (positivity rates of 0-0.13% for educators/staff and 0-0.21% for students, consistent with overall positivity rates of 0.1% in the state-wide pooled screening program).^47^ With an average pooled testing cost of $15.61/person and the highest observed weekly positivity rate (0.16% among students and educators/staff), the cost per case identified from routine asymptomatic testing in MA schools would be approximately $9,756. These data do not provide a measure of the benefit of screening programs. In estimating those benefits and deciding whether routine testing programs provide enough benefit to justify the cost, decision-makers must consider the value of test results, both in terms of potential cases averted (which were few in this study), and also in terms of real-time, locally specific data and reassurance about the safety of in-person education provided by screening programs. Without this full assessment, the interpretation of cost/case detected is difficult.

There were several limitations inherent to our study. First, district participation was incomplete and varied weekly, leading us to rely primarily on last-reported-week data. While higher and more regular participation would improve generalizability, we are thankful for voluntary survey responses by district staff already working tirelessly to implement both virtual and in-person learning during the pandemic. Second, questions about the personnel number and hours spent offered responses only in strata (e.g., 1-5 hours; 10-15 people). Our cost estimates assumed the mid-point of these ranges, although use of upper and lower bounds did not substantially change results. Third, we assigned hourly wages for parents and volunteers based on a state-wide estimate of hourly wage;^30^ local opportunity costs may have differed. Fourth, costs reported here do not include the materials needed at school to administer tests, including disposable gloves, hand sanitizer, cleaning supplies, and personal protective equipment for staff.^36,48^

Importantly, the 21 districts in this study include a lower proportion of Black and Hispanic students, English-language learners, and economically disadvantaged students than the state-wide average. While the costs of assays will likely be similar in most settings, resources needed for implementation may differ widely, for example, the availability of parent volunteers and the time needed for outreach, education, and obtaining consent for all participating students. The Massachusetts state-supported program paid many of the costs for participating districts; other districts used privately-raised funds and/or Coronavirus Aid, Relief, and Economic Security Act/Elementary and Secondary Emergency Relief Funds funds.^49,50^ Without state or federal funding, access to screening will be inequitable.

As schools offer in-person learning in 2021-22 with fewer mitigation measures and viral variants with greater transmissibility than in 2020-21, information about the resources needed to implement CDC-recommended screening programs can inform program planning.^51^

## Supporting information

Appendix A_Survey

Appendix B_Costing Calculations

## Data Availability

All data produced in the present work are contained in the manuscript

## REFERENCES

1. CDC, CDC. Community, Work, and School. Centers for Disease Control and Prevention. Published February 11, 2020. Accessed July 21, 2021. https://www.cdc.gov/coronavirus/2019-ncov/community/schools-childcare/operation-strategy.html

2. Ciaranello A, Bell T. Using Data and Modeling to Understand the Risks of In-Person Education. JAMA Netw Open. 2021;4(3):e214619. doi:10.1001/jamanetworkopen.2021.4619

3. Naimark D, Mishra S, Barrett K, et al. Simulation-Based Estimation of SARS-CoV-2 Infections Associated With School Closures and Community-Based Nonpharmaceutical Interventions in Ontario, Canada. JAMA Netw Open. 2021;4(3):e213793. doi:10.1001/jamanetworkopen.2021.3793

4. Zimmerman KO, Akinboyo IC, Brookhart MA, et al. Incidence and Secondary Transmission of SARS-CoV-2 Infections in Schools. Pediatrics. 2021;147(4):e2020048090. doi:10.1542/peds.2020-048090

5. CDC. Guidance for COVID-19 Prevention in K-12 Schools and ECE Programs. Centers for Disease Control and Prevention. Published July 9, 2021. Accessed July 21, 2021. https://www.cdc.gov/coronavirus/2019-ncov/community/schools-childcare/k-12-guidance.html

6. Tupper P, Colijn C. COVID-19 in schools: Mitigating classroom clusters in the context of variable transmission. PLOS Comput Biol. 2021;17(7):e1009120. doi:10.1371/journal.pcbi.1009120

7. Falk A, Benda A, Falk P, Steffen S, Wallace Z, Høeg TB. COVID-19 Cases and Transmission in 17 K–12 Schools — Wood County, Wisconsin, August 31–November 29, 2020. MMWR Morb Mortal Wkly Rep. 2021;70(4):136–140. doi:10.15585/mmwr.mm7004e3

8. CDC. Healthcare Workers. Centers for Disease Control and Prevention. Published February 11, 2020. Accessed July 21, 2021. https://www.cdc.gov/coronavirus/2019-ncov/hcp/testing-overview.html

9. Rafiei Y, Mello MM. The Missing Piece — SARS-CoV-2 Testing and School Reopening. N Engl J Med. 2020;383(23):e126. doi:10.1056/NEJMp2028209

10. Doron S, Ingalls RR, Beauchamp A, et al. Weekly SARS-CoV-2 Screening of Asymptomatic Students and Staff to Guide and Evaluate Strategies for Safer in-Person Learning. Infectious Diseases (except HIV/AIDS); 2021. doi:10.1101/2021.03.20.21253976

11. Pollock NR, Berlin D, Smole SC, et al. Implementation of SARS-CoV2 Screening in K-12 Schools Using In-School Pooled Molecular Testing and Deconvolution by Rapid Antigen Test. Public and Global Health; 2021. doi:10.1101/2021.05.03.21256560

12. Bi C, Mendoza R, Cheng H-T, et al. Pooled Surveillance Testing Program for Asymptomatic SARS-CoV-2 Infections in K-12 Schools and Universities. Infectious Diseases (except HIV/AIDS); 2021. doi:10.1101/2021.02.09.21251464

13. Bilinski A, Ciaranello A, Fitzpatrick MC, et al. Asymptomatic COVID-19 Screening Tests to Facilitate Full-Time School Attendance: Model-Based Analysis of Cost and Impact. Infectious Diseases (except HIV/AIDS); 2021. doi:10.1101/2021.05.12.21257131

14. Ciaranello A, Goehringer C, Nelson SB, Ruark LJ, Pollock NR. Lessons learned from implementation of SARS-CoV-2 screening in K-12 public schools in Massachusetts. Open Forum Infect Dis. Published online June 4, 2021:ofab287. doi:10.1093/ofid/ofab287

15. Covid-19 Testing in K-12 Settings: A Playbook for Educators and Leaders. The Rockefeller Foundation. Accessed September 14, 2021. https://www.rockefellerfoundation.org/report/covid-19-testing-in-k-12-settings-a-playbook-for-educators-and-leaders/

16. Updates to DESE COVID-19 Guidance. Massachusetts Department of Elementary and Secondary Education. Published May 27, 2021. https://www.doe.mass.edu/covid19/on-desktop/covid19-guide-updates.pdf

17. CDC. Coronavirus Disease 2019 (COVID-19). Centers for Disease Control and Prevention. Published February 11, 2020. Accessed July 26, 2021. https://www.cdc.gov/coronavirus/2019-ncov/science/science-briefs/transmission_k_12_schools.html

18. Wang CJ, Bair H. Operational Considerations on the American Academy of Pediatrics Guidance for K-12 School Reentry. JAMA Pediatr. 2021;175(2):121. doi:10.1001/jamapediatrics.2020.3871

19. Du Z, Pandey A, Bai Y, et al. Comparative cost-effectiveness of SARS-CoV-2 testing strategies in the USA: a modelling study. Lancet Public Health. 2021;6(3):e184–e191. doi:10.1016/S2468-2667(21)00002-5

20. Schools Should Prioritize Reopening in Fall 2020, Especially for Grades K-5, While Weighing Risks and Benefits. National Academies of Sciences, Engineering, and Medicine. Published July 15, 2021. Accessed July 26, 2021. https://www.nationalacademies.org/news/2020/07/schools-should-prioritize-reopening-in-fall-2020-especially-for-grades-k-5-while-weighing-risks-and-benefits

21. Abbott B. Covid-19 Testing in Schools Bolsters Safety but Is Hard to Set Up, Studies Find. Wall Street Journal. https://www.wsj.com/articles/covid-19-testing-in-schools-bolsters-safety-but-is-hard-to-set-up-studies-find-11612414860. Published February 4, 2021. Accessed September 14, 2021.

22. Wellesley Education Foundation | Back-to-School Testing Program. WEF. Accessed July 26, 2021. https://www.wellesleyeducationfoundation.org/testing-collaborative

23. Pollock NR, Berlin D, Smole SC, et al. Implementation of SARS-CoV2 Screening in K-12 Schools Using In-School Pooled Molecular Testing and Deconvolution by Rapid Antigen Test. Public and Global Health; 2021. doi:10.1101/2021.05.03.21256560

24. Open and Safe Schools. Open & Safe Schools. Accessed July 26, 2021. https://www.openandsafeschools.org

25. Boehm JS. The power of parent scientists. Cell. 2021;184(9):2263–2270. doi:10.1016/j.cell.2021.03.049

26. Coronavirus/COVID-19: Pooled Testing in K-12 Schools. Accessed July 26, 2021. https://www.doe.mass.edu/covid19/pooled-testing/

27. Coronavirus/COVID-19: COVID-19 Testing Program. Accessed September 13, 2021. https://www.doe.mass.edu/covid19/testing/

28. Profiles Help -About the Data. School and District Profiles. Accessed July 28, 2021. https://profiles.doe.mass.edu/help/data.aspx?section=students#selectedpop

29. School and District Profiles. Accessed July 27, 2021. https://profiles.doe.mass.edu/

30. Massachusetts - May 2020 OEWS State Occupational Employment and Wage Estimates. Accessed July 27, 2021. https://www.bls.gov/oes/current/oes_ma.htm

31. Employer Costs for Employee Compensation Summary. Accessed October 27, 2021. https://www.bls.gov/news.release/ecec.nr0.htm

32. Occupational Employment and Wage Statistics Home Page. Accessed October 27, 2021. https://www.bls.gov/oes/home.htm

33. Abdalhamid B, Bilder CR, Garrett JL, Iwen PC. Cost Effectiveness of Sample Pooling to Test for SARS-CoV-2. J Infect Dev Ctries. 2020;14(10):1136–1137. doi:10.3855/jidc.13935

34. Meiselbach MK, Bai G, Anderson GF. Charges of COVID-19 Diagnostic Testing and Antibody Testing Across Facility Types and States. J Gen Intern Med. Published online September 15, 2020. doi:10.1007/s11606-020-06198-y

35. Simas AM, Crott JW, Sedore C, et al. Pooling for SARS-CoV2 Surveillance: Validation and Strategy for Implementation in K-12 Schools. Published online December 16, 2020:2020.12.16.20248353. Accessed September 2, 2021. https://www.medrxiv.org/content/10.1101/2020.12.16.20248353v1

36. Rice KL. Estimated Resource Costs for Implementation of CDC’s Recommended COVID-19 Mitigation Strategies in Pre-Kindergarten through Grade 12 Public Schools — United States, 2020–21 School Year. MMWR Morb Mortal Wkly Rep. 2020;69. doi:10.15585/mmwr.mm6950e1

37. Baker-Polito Administration Announces Pooled Testing Initiative for Massachusetts Schools, Districts | Mass.gov. Accessed July 27, 2021. https://www.mass.gov/news/baker-polito-administration-announces-pooled-testing-initiative-for-massachusetts-schools

38. MA students return to school to new COVID testing programs. Boston 25 News. Accessed September 13, 2021. https://www.boston25news.com/news/health/ma-students-return-school-new-covid-testing-programs/4QBRJK57O5HUZHS6LEQIH4OW4Y/

39. Furfaro H. A handful of Washington schools are rapid testing staff and students for COVID-19. Is it working? The Seattle Times. Published February 8, 2021. https://www.seattletimes.com/education-lab/a-handful-of-washington-schools-are-rapid-testing-staff-and-students-for-covid-19-is-it-working/

40. Washington State School-Based COVID-19 Rapid Testing Program. Seattle Children’s Hospital. Accessed September 13, 2021. https://www.seattlechildrens.org/research/centers-programs/science-education-department/school-covid-testing/

41. Standard-Examiner EA. Davis School District making second attempt to employ “Test to Stay” at Davis High School. Standard-Examiner. Accessed July 27, 2021. https://www.standard.net/news/education/davis-school-district-making-second-attempt-to-employ-test-to-stay-at-davis-high-school/article_aa93f2c8-dced-5a06-9235-b839412178bf.html

42. Dashboard - Davis School District. Accessed September 13, 2021. https://www.davis.k12.ut.us/departments/risk-management/covid19/dashboard

43. Booth D. Michigan health department launches program offering weekly COVID testing to educators. WDIV. Published February 3, 2021. Accessed July 27, 2021. https://www.clickondetroit.com/news/michigan/2021/02/03/michigan-health-department-launches-program-offering-weekly-covid-testing-to-educators/

44. Coronavirus - MDHHS to provide COVID tests to educators to keep staff, students and community safe as schools offer in-person learning. Accessed September 13, 2021. https://www.michigan.gov/coronavirus/0,9753,7-406-98158-551193--,00.html

45. Faherty LJ, Master BK, Steiner ED, et al. COVID-19 Testing in K–12 Schools: Insights from Early Adopters. RAND Corporation; 2021. Accessed September 1, 2021. https://www.rand.org/pubs/research_reports/RRA1103-1.html

46. Governor Cuomo Announces New Testing Initiatives to Improve COVID-19 Detection & Control Across New York State. Accessed July 27, 2021. https://www.governor.ny.gov/news/governor-cuomo-announces-new-testing-initiatives-improve-covid-19-detection-control-across-new

47. Coronavirus/COVID-19: Positive COVID-19 Cases in Schools. Accessed September 1, 2021. https://www.doe.mass.edu/covid19/positive-cases/

48. Binnicker MJ. Challenges and Controversies to Testing for COVID-19. Kraft CS, ed. J Clin Microbiol. 2020;58(11). doi:10.1128/JCM.01695-20

49. Elementary and Secondary School Emergency Relief Fund Tracker. Accessed July 28, 2021. https://www.ncsl.org/ncsl-in-dc/standing-committees/education/cares-act-elementary-and-secondary-school-emergency-relief-fund-tracker.aspx

50. Division N. Biden Administration to Invest More Than $12 Billion to Expand COVID-19 Testing. HHS.gov. Published March 17, 2021. Accessed July 29, 2021. https://www.hhs.gov/about/news/2021/03/17/biden-administration-invest-more-than-12-billion-expand-covid-19-testing.html

51. Lam-Hine T. Outbreak Associated with SARS-CoV-2 B.1.617.2 (Delta) Variant in an Elementary School — Marin County, California, May–June 2021. MMWR Morb Mortal Wkly Rep. 2021;70. doi:10.15585/mmwr.mm7035e2

