## Appendix A_Survey for "Resources Required for Implementation of SARS-CoV-2 Screening in Massachusetts K-12 Public Schools in Winter/Spring 2021"

**Appendix A: Massachusetts Safer Teachers Safer Students (STSS) SARS-CoV-2 Screening: K-12 Public School Survey**

The data collected from this survey will assist other K-12 public schools and policymakers in supporting SARS-CoV-2 screening in two ways:

First, information about learning models and screening strategy will be shared on a public STSS Data Dashboard for each participating district. The dashboard will provide: an updated snapshot of screening practices other districts are taking; an FAQ page to help newcomers to screening to get off the ground running; and, a source of information for communities and policy makers deciding when to open some level of in-person learning by giving them insight into what other districts are doing and under what circumstances. Questions in this survey that will supply information for the public dashboard are noted at the start of each section.

Second, data from this survey will be used for a research study to evaluate the feasibility, acceptability, affordability, and impact of routine SARS-CoV-2 screening among asymptomatic K-12 students and educators/staff. Data collection using this survey was approved by the Mass General Brigham Institutional Review Board for Massachusetts public schools. The goal of this research is to identify barriers, facilitators, and best practices for SARS-CoV-2 screening in schools, in order to understand the potential role that screening programs may play in facilitating safe in-person learning for public school districts with widely varying access to resources. All data for the research study will be deidentified and combined with data from other districts; no individual districts or towns/municipalities will be identified. If you have any questions, please reach out to Andrea Ciaranello, MD, MPH at.

Please complete this survey weekly even if your screening program tests people more or less frequently than this. If nothing has changed since the last screening event, there are options to skip each relevant section.

**Section A: Basic Information**
A1. Email
A2. Your name
A3. District name
A4. Your role in this district (e.g. superintendent, school nurse, project manager, volunteer)
A4. Please provide the date for the MONDAY of the week that you are reporting on here

**Section B: Current district-level SARS-CoV-2 screening questions (summary information from this section to be displayed on the STSS data dashboard)**

B1. Q1. Testing/screening population - To whom are you currently OFFERING screening for SARS-CoV-2? (check all that apply)
NO screening currently being conducted
All Educators (in student-facing roles)
Some or all staff (e.g. custodial and food services, administration, others)
Some or all students
Random, or non-random, sub-samples of students or educators/staff (describe below)

B2. If you indicated in Q1 that you are screening random- or non-random sub-samples, who and how are you screening (e.g., a 10% random sample of students, or staff participating in specific activities, etc.)?

B3. For which school level(s) are you conducting SARS-CoV-2 screenings for EDUCATORS/STAFF? (check all that apply)
N/A, none
Elementary
Middle
High
ONLY High needs, ELL educators etc (spanning multiple grades)
District-wide (e.g. custodial, transportation etc)

B4. For which school level(s) are you conducting SARS-CoV-2 screenings for STUDENTS? (check all that apply)
N/A, none
Elementary
Middle
High
ONLY High needs (spanning multiple grades)

**Section C: Current district learning model (summary information from this section to be displayed on the STSS data dashboard)**

C1. Current Learning Model (check all that apply)
Remote learning (some or all grades)
Temporary remote learning - schools or classes with positive cases identified
Hybrid – some, or all, students in person some days (1,2,3 full or half days) each week (select relevant grades in question 5)
Full in person – students at specific grade levels are permitted in school 4-5 full days/week (select relevant grades in question 5)
High needs students are learning in person (spanning multiple grades)

C2. If hybrid, please select current hybrid learning model (check all that apply)
No students currently in hybrid learning
2 full days in class, 3 days remote per week
2 half days in class per week
1 full week in class with followed by 1 full week remote
4-5 half days in class per week
3-4 full days — 3-4 full days per week
Other (please describe other hybrid type below)

C3. If “other”, provide more details about the hybrid model that your district is currently pursuing:

C4. At which grade levels are students in schools for at least some in-person learning (check all that apply)
None
All grades

Pre-K
K-12 (individual select)
High needs students only (spanning multiple grades)

C5. For students attending school in person, are any classrooms laid out with LESS than 6 ft distance (nose to nose) between individuals?
Yes
No

C6. If yes to Q6. please briefly describe the distancing strategy in classrooms with LESS than 6 ft between students:

**Section D: Screening strategy: EDUCATORS/STAFF (summary information from this section to be displayed on the STSS data dashboard)**

D1. Current screening frequency
1x week screening
2x week screening
3x week screening
Once ~every two weeks
Once per month (~every 4 weeks)
Baseline screening (i.e. one-time initial screening, or one-time return-from-vacation baseline screening)

D2. Which of the following educators/staff are currently being screened? (check all that apply)
Educators
Administration
Transportation
Custodial
Food service
Athletics staff
Substitute staff
Other Professional staff (Nurses, PT, OT, etc)
Afterschool programming staff
Others - specify below
Don't know

D3. If you selected Others in Q2, please specify what role(s)

D4. Screening location (check all that apply)
At school
At centralized location
At home

D5. What sample collection process does your school district use to screen educators/staff for SARS-CoV-2? (district level only)
Observed self-collected
Unobserved self-collected
Collected by healthcare or school provider (RN, EMT, MD, etc)

D6. What specimen type is being collected for screening of Educators/Staff? (check all that apply)
Saliva
Anterior nares (front or mid-nose) swab
Nasopharyngeal (deep back of nose) swab
Don't know (please find out for next time filling in the survey)

D7. What assay type is being used for screening of Educators/Staff? (check all that apply)
PCR (sent to lab)
Rapid test (processed on site)
Don't know (please find out for next time filling in the survey)

D8. If assay type for Educators/Staff is PCR:
Individual PCR
Pooled PCR (please also complete section on pooling strategy)
N/A

D9. If assay type for Educators/Staff is rapid test, what type:
Rapid antigen (e.g. Abbott BinaxNOW, BD Veritor, etc)
Rapid molecular (e.g. Abbott ID NOW)
N/A

D10. For Educators/Staff currently being tested, what grade levels are they working with? (check all that apply)
High needs students ONLY (spanning multiple grades)
All grades
Pre-K
K-12 (individually select)
District-wide (e.g. custodial, transportation etc)
Don't know

D11. How are NEGATIVE results returned to Educators/Staff (check all that apply)?
No reporting for negative results
Educators/staff have access to individual log-in to a dashboard for results
Results/reports emailed and/or texted
Don't know

D12. How are POSITIVE results returned to Educators/Staff (check all that apply)?
Educators/staff have access to individual log-in to a dashboard for results
Results/reports emailed and/or texted
They are called in person

D13. If contacted by phone, who makes the calls to Educators/Staff with positive results?
School nurse
Other school staff member (e.g. superintendent, project manager)
Board of Health
Third party vendor
Don't know

**Section E: Screening strategy: STUDENTS**
E1. Current Screening Frequency: STUDENTS
1x week
2x week
Once every 2 weeks
Once every 4 weeks — ~once per month
Baseline screening — i.e. one-time initial screening, or one-time return-from-vacation baseline

E2. Screening location for Students (check all that apply)
At school
At centralized location
At home

E3. What collection process does your school district use to screen Students for SARS-CoV-2? (check all that apply)
Observed self-collected
Unobserved self-collected
Parent-collected
Collected by healthcare or school provider (RN, EMT, MD, etc)
Don't know (please find out for next time filling out the survey)

E4. At what GRADE threshold do you allow self-collection of samples by Students?

E5. What specimen type is being collected for screening of Students?
Saliva
Anterior nares (front or mid-nose) swab
Nasopharyngeal (deep back of nose) swab
Don't know

E6. What assay type is being collected for screening of Students?
PCR (sent to lab)
Rapid test (processed on site)

E7. If assay type is PCR for Student testing:
Individual PCR
Pooled PCR (please also complete section on pooling strategy)
N/A

E8. If assay type is rapid test for Student testing:
Rapid antigen (e.g. Abbott BinaxNOW, Veritor BD, etc)
Rapid molecular (e.g. Abbott ID NOW)
N/A

E9. What grade levels of students are currently being offered SARS-CoV-2 asymptomatic screening? (check all that apply)
All grades
Pre-K

K-12
High needs ONLY (spanning multiple grades)

E10. How are NEGATIVE results returned to students/parents? (Check all that apply)
No reporting for negative results
Students/parents have access to an individual log-in dashboard for results
Results/reports are emailed and/or texted

E11. How are POSITIVE results returned to students/parents? (check all that apply)
Students/parents have access to an individual log-in dashboard for results
Results/reports are emailed and/or texted
Students/parents are called in person

E12. If contacted by phone, who makes the calls to students/parents with positive results? (check all that apply)
School nurse
Other school staff member (e.g. superintendent, project manager etc)
Third party vendor
Board of Health
Don't know

**Section F and G: Pooled screening: EDUCATORS/STAFF ONLY and STUDENTS ONLY**
F/G1. Are you specifically creating pools with Educator/Staff ONLY or STUDENTS ONLY (Answer NO if students are included in the pools by design, or if your pool design is random)?
Yes
No

F/G2. If YES to Q1, what is the average size of the EDUCATORS/STAFF ONLY or STUDENT ONLY pools?
8
10
12
20
24

F/G3. At what site are the samples from Educators/Staff or Students physically pooled together?
On site in a single tube as we collect them (e.g. we pool the swabs together in a large tube)
After we send individual samples to the lab (the lab does the sample pooling)
Don't know

F/G4. What is your pooling strategy – how do you combine samples for Educators/Staff ONLY or STUDENTS ONLY in pools together?
Randomly
By classroom or cohort
By grade level
By discipline (e.g. all athletics staff)
Don't know
Other - describe below

F/G6. What is the process for returning individual results (deconvoluting, or reflex testing) from an Educator/Staff ONLY or STUDENT ONLY pool that tests positive (check all that apply)?
Lab saves individual samples, retests all and returns INDIVIDUAL results from a positive pool
Lab saves individual samples, retests in pools of 2, then tests and returns INDIVIDUAL results from the positive pool of 2
Everyone in positive pools needs rescreening (we collect a new sample for the SAME type of test)
Everyone in positive pool needs rescreening (we collect a new sample for a RAPID test)
Other - describe below

F/G8. If reflex testing/re-screening individuals from an Educator/Staff ONLY or STUDENT ONLY positive pool, what is the test type used for this second test (Check all that apply)?
N/A (lab saves and retests the original samples for us)
Saliva swab for PCR
Anterior nares swab for PCR
Nasopharyngeal swab for PCR
Anterior nares swab for rapid test (e.g. Abbott BinaxNOW)

G8. What is your pooling strategy for students (or student/staff mixed pools) – how do you pool samples together?
Randomly
By classroom
By grade level
Don't know
Other - describe below

**Section H: Screening results: EDUCATORS/STAFF and STUDENTS**
H1. APPROXIMATE number of Educators/Staff who were OFFERED (or were ELIGIBLE for) screening tests during the prior week (Monday-Sunday): [NOTE - although this number is used to estimate participation rates, we appreciate that it may be a 'best estimate' & recommend you try for best consistency in reporting weekly]

H2. Number of Educators/Staff ACTUALLY screened during the prior week (Monday-Sunday):

H3. If using pooled testing for Educators/Staff, number of positive POOLS with Staff/Educators that identified last week (Monday-Sunday) that included an Educator &/or Staff member (use N/A if not applicable):

H4. Number of INDIVIDUAL Educators/Staff that were detected as positive from screening the prior week (Monday-Sunday):

H5. At what school level(s) were your positive Educator/Staff cases identified through the screening program: (check all that apply)
Elementary
Middle
High
District-wide (e.g. custodial, transportation etc)
N/A

H6. Time from shipment to return of 90% (i.e. the vast majority) of INITIAL results for Educators/Staff last week (Monday to Sunday)?
<12 hours
<24 hours
<48 hours
<72 hours

H7. If pooled testing was used for Educators/Staff, how much additional time was needed for retesting (reflex testing) of individuals in positive pools (testing time but not including contact tracing time)?
N/A
Additional <6 hours
Additional ~12 hours
Additional ~24 hours
Additional ~48 hours

H8. If there were any unusual events last week that caused a delay in return of results for Staff/Educator pools, please describe below:

H9. Approximately what proportion of your educators/staff have been vaccinated against SARS-CoV-2 (as of this reporting week)?

**Section I: Effect of Screening Results on Learning Model**
I1. Did any of your schools, or classes, switch to temporary remote learning last week due to positive cases in your school community?
Not applicable (no positive cases identified)
No (even though positive cases were identified)
1 or more Elementary schools or classrooms
1 or more Middle schools or classrooms
1 or more High schools or classrooms

I2. Were these positive cases identified via the asymptomatic screening program or through symptomatic testing of individual(s) displaying covid-like illness? (check all that apply)
Not applicable
Asymptomatic screening program
Tested after displaying symptoms
Tested as a 'close contact' of a positive case

I3. Were the positive cases that triggered your switch to remote learning detected among educators/staff or students?
Not applicable
Educators/staff
Students
Both educators/staff and students
Not disclosed

**Section J: Vendors and funding**
J1. What vendor does your district use to initially screen EDUCATORS/STAFF?
Mirimus

CIC Health
Ginkgo (Concentric)
JCM Analytics
Project Beacon

J2. What vendor does your district use to initially screen STUDENTS?
Mirimus
CIC Health
Ginkgo (Concentric)
JCM Analytics
Project Beacon

J3. What vendor(s) does your district use for reflex testing (retesting of individuals in a positive pool)? (Check all that apply if using more than one, or a different approach for Students vs Educators/Staff)
N/A. We are NOT doing pooled testing
Mirimus
CIC Health
Ginkgo (Concentric)
JCM Analytics
Project Beacon
BinaxNOW

J4. Who is the ordering/notifying medical provider for the screening program?
District physician
Board of Health physician
Town or City physician
Physician provided by vendor (e.g. PWN Health, etc)
Other (please describe below)
Don't know

J5. What is the approximate cost per person tested to screen EDUCATORS/STAFF for SARS-CoV-2? (Include only materials and lab analysis, do not include estimated staff time) Specify or write in "N/A":

J6. What is the approximate cost per person tested to screen STUDENTS for SARS-CoV-2? (Include only materials and lab analysis, do not include estimated staff time) Specify or write in "N/A":

J7. How is your district covering the cost of the screening tests? Check all that apply.
City funding
CARES/ARP funding
Parent fees
Private Foundations
School district budget
Stimulus funding
Other (describe below)

J8. Who is participating in the implementation of the SARS-CoV-2 screening process(es)? (Check all that apply)
School nurses at the district level

School nurses at each school
Paid project manager
Volunteer project manager
District physician or medical advisory team
District HHS Department
Town or city staff
Educators/school staff
Parent volunteers
Other
Don't know

J9. How many people in total are typically involved in the implementation of the screening program over the course of a full week (Monday to Sunday) EXCLUDING contact tracing?
1-5 people
5-10 people
10-15 people
15-20 people
20-25 people
>25 people

J10. How many TOTAL hours, COMBINED, are spent by all staff to implement the program in a typical week (Monday to Sunday) (e.g., 5 people each worked ~5 hours this week = 25 hours)?
5-10 hours/week
10-15 hours/week
15-20 hours/week
20-25 hours/week
>25 hours/week
>50 hours/week
Don't know (please find out by the next time you fill in the survey)

Archive:::::

**Section H: Pooled screening: STUDENTS & MIXED POOLS**

H1. Are you creating mixed pools that might include Students AND Educators/Staff?
Yes - we deliberately create Student/Staff mixed pools (answer next question)
Yes - but our pools are random (student/staff numbers vary in each pool)
No - all of our pools are STUDENTS ONLY

H2. If you SPECIFICALLY design/create Student/Staff mixed pools, how many staff are you including in each mixed pool (answer N/A if your pools are random)?

Q3. What is the TOTAL POOL SIZE for Student only, or Mixed student/staff pools?
8
10
12
20
24

Q4. At what site are the samples for Student &/or Mixed pools physically pooled together?
On site in a single tube as we collect them (e.g. we pool swabs together in a large tube)
After we send individual samples to the lab (the lab does the sample pooling)
Don't know

Q5. What is the process for returning individual results (deconvoluting, or reflex testing) from a Student only &/or Mixed pool that tests positive (check all that apply)?
Lab saves individual samples, retests all and returns INDIVIDUAL results from a positive pool
Lab saves individual samples, retests in pools of 2, then tests and returns INDIVIDUAL results from the positive pool of 2
Everyone in positive pools needs rescreening (we collect a new sample for SAME type of test)
Everyone in positive pool needs rescreening (we collect a new sample for a RAPID test)
Other - describe below

Q6. If “other” for deconvoluting results, please describe:

Q7. If reflex testing/rescreening individuals from a Student only &/or mixed positive pool, what is the test type used for this second test (Check all that apply)?
N/A (lab saves and retests the original samples for us)
Saliva swab for PCR
Anterior nares swab for PCR
Nasopharyngeal swab for PCR
Anterior nares swab for rapid test (e.g. Abbott BinaxNOW)
