## Appendix B_Costing Calculations for "Resources Required for Implementation of SARS-CoV-2 Screening in Massachusetts K-12 Public Schools in Winter/Spring 2021"

| | Total personnel reported | Total hours reported | Role | Assumed number of people in each role (sum to B) | Relative weight (sum to 1) | Weighted # hours for each role (sum to C) | Hourly wage for each role | Weekly cost for each role | Per-person assay cost (students) | Total weekly personnel cost | # screened per week (students) | Total assay + personnel weekly cost |
|---|---|---|---|---|---|---|---|---|---|---|---|---|
| 1 | 18 | 13 | School nurses at each school | 8.5 | 0.47 | 6.14 | $66.84 | $410 | | | | |
| | 18 | 13 | School nurses at each district | 8.5 | 0.47 | 6.14 | $66.84 | $410 | | | | |
| | 18 | 13 | Paid project managers | 1 | 0.06 | 0.72 | $42.52 | $31 | $5.00 | **$851** | 101.00 | $1,356.36 |
| 2 | 13 | 23 | School nurses at each school | 3.666667 | 0.28 | 6.49 | $66.84 | $434 | | | | |
| | 13 | 23 | School nurses at each district | 3.666667 | 0.28 | 6.49 | $58.42 | $379 | | | | |
| | 13 | 23 | Paid project managers | 1 | 0.08 | 1.77 | $42.52 | $75 | | | | |
| | 13 | 23 | District HHS department | 1 | 0.08 | 1.77 | $22.26 | $39 | | | | |
| | 13 | 23 | Educators/school staff | 3.666667 | 0.28 | 6.49 | $58.42 | $379 | $5.00 | **$1,306** | 28.00 | $1,446.22 |
| 3 | 3 | 100 | School nurses at each school | 0.6 | 0.20 | 20.00 | $66.84 | $1,337 | | | | |
| | 3 | 100 | School nurses at each district | 0.6 | 0.20 | 20.00 | $66.84 | $1,337 | | | | |
| | 3 | 100 | Educators/school staff | 0.6 | 0.20 | 20.00 | $58.42 | $1,168 | | | | |
| | 3 | 100 | Parent volunteer | 0.6 | 0.20 | 20.00 | $25.75 | $515 | | | | |
| | 3 | 100 | Other | 0.6 | 0.20 | 20.00 | $31.93 | $639 | $5.00 | **$4,996** | 624.00 | $8,115.68 |
| 4 | 18 | 18 | School nurses at each school | 4.50 | 0.25 | 4.50 | $66.84 | $301 | | | | |
| | 18 | 18 | School nurses at each district | 4.50 | 0.25 | 4.50 | $66.84 | $301 | | | | |
| | 18 | 18 | Educators/school staff | 4.50 | 0.25 | 4.50 | $58.42 | $263 | | | | |

| | | | | | | | | | | | | |
|---|---|---|---|---|---|---|---|---|---|---|---|---|
| | 18 | 18 | Other | 4.50 | 0.25 | 4.50 | $31.93 | $144 | $15.00 | **$1,008** | 1295.00 | $20,433.15 |
| 5 | 8 | 18 | School nurses at each school | 7.5 | 0.94 | 16.88 | $66.84 | $1,128 | $10.00 | **$1,128** | 874.00 | $9,867.93 |
| 6 | 23 | 38 | District HHS department | 1 | 0.04 | 1.65 | $22.26 | $37 | | | | |
| | 23 | 38 | Superintendent | 1 | 0.04 | 1.65 | $75.29 | $124 | | | | |
| | 23 | 38 | School nurses at each school | 12 | 0.52 | 19.83 | $66.84 | $1,325 | | | | |
| | 23 | 38 | Volunteer project managers | 3 | 0.13 | 4.96 | $42.52 | $211 | | | | |
| | 23 | 38 | Other | 3 | 0.13 | 4.96 | $31.93 | $158 | $25.00 | **$1,855** | 770.00 | $21,105.37 |
| 7 | 3 | 100 | School nurses at each school | 1.00 | 0.33 | 33.33 | $66.84 | $2,228 | | | | |
| | 3 | 100 | Educators/school staff | 1.00 | 0.33 | 33.33 | $58.42 | $1,947 | | | | |
| | 3 | 100 | Other | 1.00 | 0.33 | 33.33 | $31.93 | $1,064 | $5.00 | **$5,240** | 344.00 | $6,959.80 |
| 8 | 13 | 100 | School nurses at each school | 6.00 | 0.46 | 46.15 | $66.84 | $3,085 | | | | |
| | 13 | 100 | Paid project managers | 1.00 | 0.08 | 7.69 | $42.52 | $327 | | | | |
| | 13 | 100 | Town or city staff | 6.00 | 0.46 | 46.15 | $21.27 | $982 | $5.00 | **$4,394** | 772.00 | $8,253.71 |
| 9 | 38 | 23 | School nurses at each school | 9.25 | 0.24 | 5.60 | $66.84 | $374 | | | | |
| | 38 | 23 | School nurses at each district | 9.25 | 0.24 | 5.60 | $66.84 | $374 | | | | |
| | 38 | 23 | Paid project managers | 1.00 | 0.03 | 0.61 | $42.52 | $26 | | | | |
| | 38 | 23 | Town or city staff | 9.25 | 0.24 | 5.60 | $21.27 | $119 | | | | |
| | 38 | 23 | Educators/school staff | 9.25 | 0.24 | 5.60 | $58.42 | $327 | $20.00 | **$1,220** | 747.00 | $16,160.35 |
| 10 | 38 | 100 | School nurses at each school | 12.67 | 0.33 | 33.33 | $66.84 | $2,228 | | | | |
| | 38 | 100 | School nurses at each district | 12.67 | 0.33 | 33.33 | $66.84 | $2,228 | | | | |
| | 38 | 100 | Educators/school staff | 12.67 | 0.33 | 33.33 | $58.42 | $1,947 | $15.00 | **$6,403** | 2557.00 | $44,758.47 |

| | | | | | | | | | | | | |
|---|---|---|---|---|---|---|---|---|---|---|---|---|
| 11 | 18 | 100 | School nurses at each school | 5.67 | 0.31 | 31.48 | $66.84 | $2,104 | | | | |
| | 18 | 100 | School nurses at each district | 5.67 | 0.31 | 31.48 | $66.84 | $2,104 | | | | |
| | 18 | 100 | District physician or medical advisory team | 1.00 | 0.06 | 5.56 | $78.69 | $437 | | | | |
| | 18 | 100 | Town or city staff | 5.67 | 0.31 | 31.48 | $21.27 | $670 | $5.00 | **$5,315** | 1575.00 | $13,190.24 |
| 12 | 8 | 38 | School nurses at each school | 2.67 | 0.33 | 12.67 | $66.84 | $847 | | | | |
| | 8 | 38 | School nurses at each district | 2.67 | 0.33 | 12.67 | $66.84 | $847 | | | | |
| | 8 | 38 | Educators/school staff | 2.67 | 0.33 | 12.67 | $58.42 | $740 | $5.00 | **$2,433** | 553.00 | $5,198.32 |
| 13 | 100 | 8 | School nurses at each school | 50.00 | 2.00 | 16.00 | $66.84 | $1,069 | | | | |
| | 100 | 8 | Educators/school staff | 50.00 | 2.00 | 16.00 | $58.42 | $935 | $5.00 | **$2,004** | 775.00 | $5,879.22 |
| 14 | 8 | 38 | School nurses at each school | 3.50 | 0.44 | 16.63 | $66.84 | $1,111 | | | | |
| | 8 | 38 | School nurses at each district | 3.50 | 0.44 | 16.63 | $66.84 | $1,111 | | | | |
| | 8 | 38 | Paid project managers | 2.00 | 0.25 | 9.50 | $42.52 | $404 | $11.00 | **$2,626** | 1400.00 | $18,026.38 |
| 15 | 3 | 38 | School nurses at each school | 2.5 | 0.83 | 58.42 | $66.84 | $3,905.03 | 6 | **$3,905.03** | 185.00 | $5,015.03 |
| 16 | 13 | 38 | School nurses at each school | 4.33 | 0.33 | 12.67 | $66.84 | $847 | | | | |
| | 13 | 38 | School nurses at each district | 4.33 | 0.33 | 12.67 | $66.84 | $847 | | | | |
| | 13 | 38 | Paid project managers | 4.33 | 0.33 | 12.67 | $42.52 | $539 | $15.00 | **$2,232** | 1056.00 | $18,071.87 |
| 17 | 8 | 8 | School nurses at each district | 4.00 | 0.50 | 4.00 | $66.84 | $267 | | | | |
| | 8 | 8 | Parent volunteer | 4.00 | 0.50 | 4.00 | $25.75 | $103 | $5.00 | **$370** | 750.00 | $4,120.36 |
| 18 | 23 | 23 | School nurses at each school | 7.67 | 0.33 | 7.67 | $66.84 | $512 | | | | |

| | | | | | | | | | | | | |
|---|---|---|---|---|---|---|---|---|---|---|---|---|
| | 23 | 23 | School nurses at each district | 7.67 | 0.33 | 7.67 | $66.84 | $512 | | | | |
| | 23 | 23 | Educators/school staff | 7.67 | 0.33 | 7.67 | $58.42 | $448 | $12.00 | **$1,473** | 1083.00 | $14,468.80 |
| 19 | 8 | 38 | District HHS Department | 1 | 0.13 | 4.75 | $22.26 | $105.74 | | | | |
| | 8 | 38 | School nurses at each school | 6 | 0.75 | 28.5 | $66.84 | $1,904.94 | | | | |
| | 8 | 38 | District physician or medical advisory team | 1 | 0.13 | 4.75 | $78.69 | $373.78 | 6 | **$2,384.45** | 913 | $7862.45 |
| 20 | 38 | 100 | School nurses at each school | 7.00 | 0.18 | 18.42 | $66.84 | $1,231 | | | | |
| | 38 | 100 | School nurses at each district | 7.00 | 0.18 | 18.42 | $66.84 | $1,231 | | | | |
| | 38 | 100 | Superintendent | 1.00 | 0.03 | 2.63 | $75.29 | $198 | | | | |
| | 38 | 100 | Other | 22.00 | 0.58 | 57.89 | $31.93 | $1,849 | $13.50 | **$4,509** | 1190.00 | $20,574.25 |
| 21 | 18 | 38 | School nurses at each school | 5.83 | 0.32 | 12.31 | $66.84 | $823 | | | | |
| | 18 | 38 | School nurses at each district | 5.83 | 0.32 | 12.31 | $66.84 | $823 | | | | |
| | 18 | 38 | Educators/school staff | 5.83 | 0.32 | 12.31 | $58.42 | $719 | $13.81 | **$2,364** | 2907.00 | $42,510.05 |
